# Longitudinal Changes in Lifestyle Behaviors and Cardiovascular Health During the Transition to Fatherhood: The Dad Bod Study Rationale and Design

**DOI:** 10.1101/2024.09.26.24314459

**Authors:** Matthew J. Landry, Jocelyn P. Pineda, Jaylen M. Lee, Michael A. Hoyt, Karen L. Edwards, Karen L. Lindsay, Christopher D. Gardner, Nathan D. Wong

**Affiliations:** Department of Population Health and Disease Prevention, Joe C. Wen School of Population & Public Health, University of California, Irvine; Irvine, California, USA; Biostatistics, Epidemiology & Research Design Unit, Institute for Clinical and Translational Sciences, University of California, Irvine; Irvine, California, USA; Department of Epidemiology & Biostatistics, Joe C. Wen School of Population & Public Health, University of California, Irvine; Irvine, California, USA; Department of Pediatrics, School of Medicine, University of California, Irvine; Irvine, California, USA; Susan Samueli Integrative Health Institute, Susan and Henry Samueli College of Health Sciences, University of California, Irvine; Irvine, California, USA; Stanford Prevention Research Center, School of Medicine, Stanford University; Palo Alto, California, USA; Heart Disease Prevention Program, Mary and Steve Wen Cardiovascular Division, School of Medicine, University of California, Irvine; Irvine, California, USA

**Keywords:** men’s health, cardiovascular health, paternal, fatherhood

## Abstract

**Background:** Despite the importance of the transition to fatherhood as a critical life stage among young adult men, much remains unknown about the factors predictive of ideal cardiovascular health (CVH) and how CVH is impacted as young men face new roles and responsibilities associated with fatherhood.

**Methods:** To address this gap, the Dad Bod Study is a prospective, longitudinal and observational study designed to examine how fatherhood affects young men’s CVH. A total of 125, first-time prospective fathers (men, 19-39 years) will be enrolled and followed over 1.5 years. Metrics of the American Heart Association’s “Life’s Essential 8” as well as demographic, social, and psychosocial factors will be collected at four time points ((baseline (during the pregnant partner’s 2nd trimester) 1-month postpartum, 6-months postpartum, and 1-year postpartum). The primary aims are to measure predictors of CVH among first-time fathers and describe longitudinal changes in CVH. A secondary aim is to identify best practices for recruitment, retention, and remote data collection in this population.

**Summary:** The Dad Bod Study offers a novel examination of CVH among first-time fathers, exploring how new paternal roles and responsibilities impact cardiovascular health. Findings may provide key insights into critical CVH behaviors and risk factors to monitor, preserve, and improve as young men transition to fatherhood.

## Introduction

Cardiovascular disease (CVD) is a leading cause of death among men in the United States, despite >80% of all cardiovascular events being preventable through healthy lifestyles and management of known CVD risk factors^1,2^. Maintaining health promoting behaviors and avoiding the development of adverse risk behaviors earlier in life (primordial prevention) can improve cardiovascular health (CVH) and reduce the future burden of CVD^3–5^.

Young adulthood encompasses an age range between 18 and 39 years old^6^. During this life stage many critical decisions and goal pursuits occur that can have enduring ramifications – young adults often complete their education, enter the workforce, establish social networks and romantic relationships, and potentially start a family. Recent trends indicate a steady or slightly increasing incidence of CVD in young adults contrasting with the generally decreasing rates observed in middle-aged and older adults^2,7^. Exacerbating this trend, recent nationally representative data have shown that CVH in young adults has not improved in the last decade^8^. These findings portend a potential future burden of CVD and reversal of decreasing rates of CVD in later developmental periods as current young adults age.

The transition to fatherhood among young men represents a major, discrete life event in which substantial biological, psychological, and social changes can occur ^9–11^ (**Figure 1**). Some men may be motivated to improve their health during the transition to fatherhood, while others are challenged to improve or maintain health as they experience disruption to their lifestyle routines^10–12^. A new father’s ability to choose a healthy lifestyle may also be influenced by social or socioeconomic and structural elements^13–15^. Further, the transition to fatherhood can significantly impact men’s health, as the stress and anxiety associated with new parental responsibilities may exacerbate or trigger various physical and mental health issues, including depression, which has been well documented among men during the perinatal and postpartum period^16,17^.

**Figure 1.**
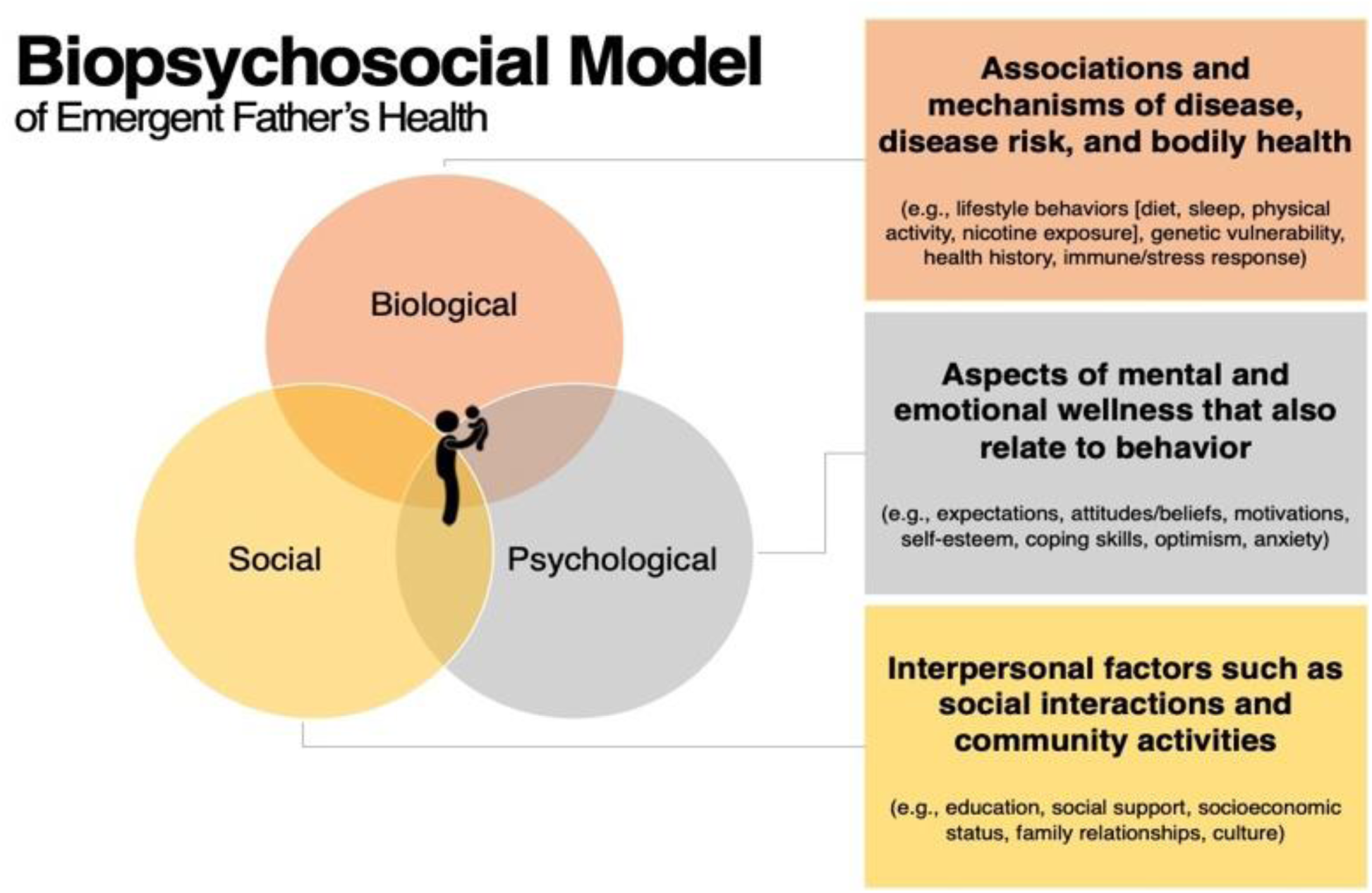
Biopsychosocial Model of the overlapping interplay of biological, psychological, and social factors that may impact emergent father’s health.

As men transition into fatherhood and throughout the postpartum period, several undesirable CVH behaviors have been observed. These include declines in physical activity and increased sedentary behavior^18,19^, reduced sleep^20–22^, poor diet quality^20,23^, and increases in body weight^19,20,22,24–27^. However, the impact of fatherhood on health behaviors is not uniformly negative. Some studies have also noted positive associations such as greater physical activity^20^ and lower rates of smoking^19^ among new fathers. Evidence also suggests that the relationship of fatherhood with CVH may also differ by age in which men enter into fatherhood and by race/ethnicity^28^. Despite the importance of the transition to fatherhood as a critical life stage among young men, much remains unknown about the factors predictive of ideal CVH and how CVH is impacted as young men face new roles and responsibilities associated with fatherhood.

The current study, titled ‘The Dad Bod Study’ was designed as a prospective, longitudinal and observational study designed to examine how fatherhood affects young men’s CVH. Specifically, the study aims to (1) examine demographic, social, and psychosocial factors that are predictive of CVH among first-time fathers, (2) examine longitudinal changes to CVH in first-time fathers using the American Heart Association’s “Life’s Essential 8” metric and (3) identify best practices for recruitment, retention, and remote data collection among first-time fathers. This article describes the design, methodology, and rationale for the Dad Bod Study.

## Methods

### Ethics Statement and Data Availability

The study was approved by the University of California, Irvine Institutional Review Board (IRB #4907, approved May 1, 2024). After study completion, the anonymized data and material will be made publicly available.

### Community Engagement Studios

In the Spring of 2024, our research team conducted a series of Community Engagement Studios (CE Studios) in partnership with the Institute for Clinical and Translational Science at the University of California, Irvine. CE Studios are a consultative model meant to provide research teams with rapid feedback from individuals representative of the population of interest on specific aspects of their existing study design before a research project is implemented^29,30^. During each CE Studio, we presented a brief overview of the research project then provided prompts regarding aspects of this project. CE Studio participants participated in a facilitated discussion and provided their opinions and recommendations. Each CE Studios focused on a different aspect of the project allowing us to gain relevant insights on recruitment methods, number of timepoints and perceptions on participant burden, data collection methods, and strategies for participant retention. Insights gathered from the CE Studios were used by the research team to refine and strengthen the methodological approach of this study described in subsequent sections.

### Participant Recruitment

When designing the study, the research team carefully considered known barriers and facilitators for recruiting and retaining male participants in longitudinal health research ^31^. Recruitment for the study is expected to begin in Fall 2024. We will recruit 125 young adult (age 19-39), male individuals. Participants will be recruited from across the United States during their pregnant partner’s 2^nd^ trimester and will be allowed to enroll in the study up until the pregnant partner’s 35^th^ week of pregnancy during the 3^rd^ trimester. First-time fathers are defined as a biological father who has not yet experienced the live birth of his own child^32^.

Multiple methods of participant recruitment will be used. Participants will be recruited using Native Health Research^33^, a participant recruitment platform that uses social media and digital advertisements to connect interested participants with research studies. Recruitment will also take place through online support groups for prospective fathers. Printed advertisements will be displayed on notice boards at various community venues in Orange County, CA, including workplaces, community group meeting locations, sports complexes, golf clubs, and local library branches. Additionally, local obstetrician-gynecologists within the university and local hospital systems will be informed about the study and provided with flyers to distribute during prenatal care visits. Interested men will complete a brief online screener, via URL link or QR code on study advertisements, to determine eligibility. As part of the screener, participants will be asked to indicate how they heard about the study during the initial enrollment process, allowing us to track and analyze the effectiveness of various recruitment methods.

### Inclusion and Exclusion Criteria

Inclusion and exclusion criteria are intended to maximize both eligibility and generalizability (**Table 1**). We will recruit only males, as the study aims seek to answer the question of how fatherhood affects men’s health. Additionally, we will exclude men aged ≥40, given our focus on management of lifestyle behaviors and cardiovascular risk factors before onset of cardiovascular disease. There will be no exclusion based on racial or ethnic status. Due to budget limitations, study materials will be provided only in English. Additionally, recognizing the diverse nature of modern families, the study adopts an inclusive approach with no eligibility restrictions on fathers’ living arrangements or planned involvement in infant care. However, this information will be collected for analytical purposes.

**Table 1.** Inclusion and Exclusion Criteria.

| Table 1. Inclusion and Exclusion Criteria |
| --- |
| Inclusion Criteria<br>1. Self-identity as male<br>2. Age ≥19 and ≤39 years<br>3. First-time biological father<br>4. Ability to read written English<br>5. Generally healthy (no confirmed diagnosis of a chronic disease (e.g., coronary heart disease, type 2 diabetes, chronic kidney disease)<br>6. Willing to participate in study data collection and assessment outlined in the study protocol over a period of about 1.5 years (baseline to 12-month follow-up after child’s birth)<br>7. Signed written informed consent |
| Exclusion Criteria<br>1. Age ≤18 or ≥40 years<br>2. Biological father of a prior child<br>3. Currently under psychiatric care or taking psychiatric medications<br>4. Prior history of disordered eating or severe body dysmorphia that may preclude being able to recall dietary intake or provide accurate measurements of their body weight from an electronic scale<br>5. Residing outside the United States or has anticipated plans to move outside of the United States within the next year<br>6. Enrolled in another competing investigational study, as per investigator discretion |

### Data Collection

Eligible participants will attend a study orientation session, conducted via video call. After being informed about the study design interested participants will provide written informed consent. Enrolled men will complete four longitudinal assessments (baseline (during the pregnant partner’s 2^nd^ trimester) 1-month postpartum, 6-months postpartum, and 1-year postpartum) over the course of the study. The baseline data collection timepoint will occur immediately after consenting to participate in the study. Time points were chosen based on feedback gathered in CE Studios and to be frequent enough to allow for comprehensive assessment of trends in CVH without being onerous on participants.

Given a general disengagement with traditional preventive care among young men^34,35^, this study utilizes novel approaches to recruit, retain, and collect all data remotely. Because of the remote nature, participants can be recruited anywhere in the United States, potentially allowing for a more diverse sample^36^. Additionally, participants will complete all aspects of the study from their home and will not have to come to any university or clinical site, therefore decreasing participant burden. Participants will be provided with instructional videos demonstrating the proper procedures for collecting at-home metrics, (e.g., blood pressure using a study-provided blood pressure cuff and at-home dried blood spot sample collection kits). Study staff will be available to offer additional assistance via email or video call, as needed. To maintain the study’s focus on the transition to fatherhood, participants will be compassionately withdrawn from the study in the event of pregnancy loss, stillbirth, or other circumstances resulting in an unsuccessful completion of the pregnancy.

### Collection of Life’s Essential 8 Cardiovascular Health Metrics

In 2022, the American Heart Association released its new iteration of quantifying CVH – “Life’s Essential 8”^37^. Life’s Essential 8 consists of 4 modifiable health behaviors four behaviors (diet, physical activity, sleep health, and smoking) and four modifiable risk factors (body weight, blood lipids, blood glucose, and blood pressure). At each of the four data collection time points, participants will complete assessments of each of the Life’s Essential 8 metrics (see **Table 2**). Each metric is designed to take <5 minutes to assess, easing participant burden. To facilitate remote data collection, participants will be mailed study-provided equipment including a digital body weight scale and blood pressure cuff at the onset of the study. Furthermore, at-home dried blood spot sample collection kits will be mailed to participant’s homes at each of the four assessment time points. Participants will be instructed to collect the sample while fasting (at least 8 hours since the prior meal) and will be encouraged to complete the sample collection at the same time of day for each of the four time points. Following the collection protocol, participants will seal their dried blood spot samples and return them for laboratory processing using the provided prepaid envelopes. Each metric will be scored on an ordinal point scoring scale of 0 to 100 points according to the American Heart Association scoring algorithm^37^. A higher score typically indicates better adherence to optimal health behaviors or risk factor management in that particular area. An overall CVH score will be calculated by summing the scores for each of the 8 metrics and dividing the total by 8, to provide a Life’s Essential 8 score ranging from 0 to 100.

**Table 2.** Life’s Essential 8 Metrics and Method of Measurement^a^.

| **Metric** | **Method of Measurement** |
| --- | --- |
| 1.Diet | Self-administrated dietary recall using diet quality photo navigation (DietID)<sup>51-53</sup>, from which a Healthy Eating Index-2020<sup>54</sup> (HEI-2020) score will be calculated. |
| 2.Physical Activity | Self-reported minutes of moderate or vigorous physical activity via International Physical Activity Questionnaire (IPAQ)<sup>55</sup>. |
| 3.Nicotine Exposure | Self-reported use of cigarettes or products containing nicotine via Behavioral Risk Factor Surveillance System Tobacco Use core questions<sup>56</sup>. |
| 4.Sleep Health | Self-reported average hours of sleep per night via Pittsburgh Sleep Quality Index (PSQI) item<sup>57</sup>. |
| 5.Body Mass Index (BMI) | Objective measurement of weight from study-provided scale (Tanita HS-302 Light Powered Digital Scale) and self-reported height. BMI calculated as kg/m<sup>2</sup>. |
| 6.Blood Lipids | Objective measurement of plasma total and HDL-cholesterol with calculation of non-HDL-cholesterol and fasting blood glucose from at-home dried blood spot sample collection<sup>58</sup> (Molecular Testing Labs). |
| 7.Blood Glucose |  |
| 8.Blood Pressure | Objective measurement of systolic and diastolic blood pressure from study-provided blood pressure cuff (Omron Bronze Upper Arm Monitor). |
| <sup>a</sup>All eight metrics will be collected at four assessment time points: BL: baseline (during the pregnant partner's 2<sup>nd</sup> trimester); 1M: 1-month postpartum; 6M: 6-months postpartum; and 12M: 12-month postpartum) |  |

### Questionnaires and Other Assessment Measures

The constructs, measures used, and time points at which questionnaire data collection will occur are outlined in **Table 3**. Some questionnaires are collected only once as they are relevant to only one phase of the study. Other measures are collected at multiple time points to observe change over time. Because the 1-month postpartum time point is anticipated to be a more onerous time for participants, questionnaire data collection is intentionally limited.

**Table 3.**
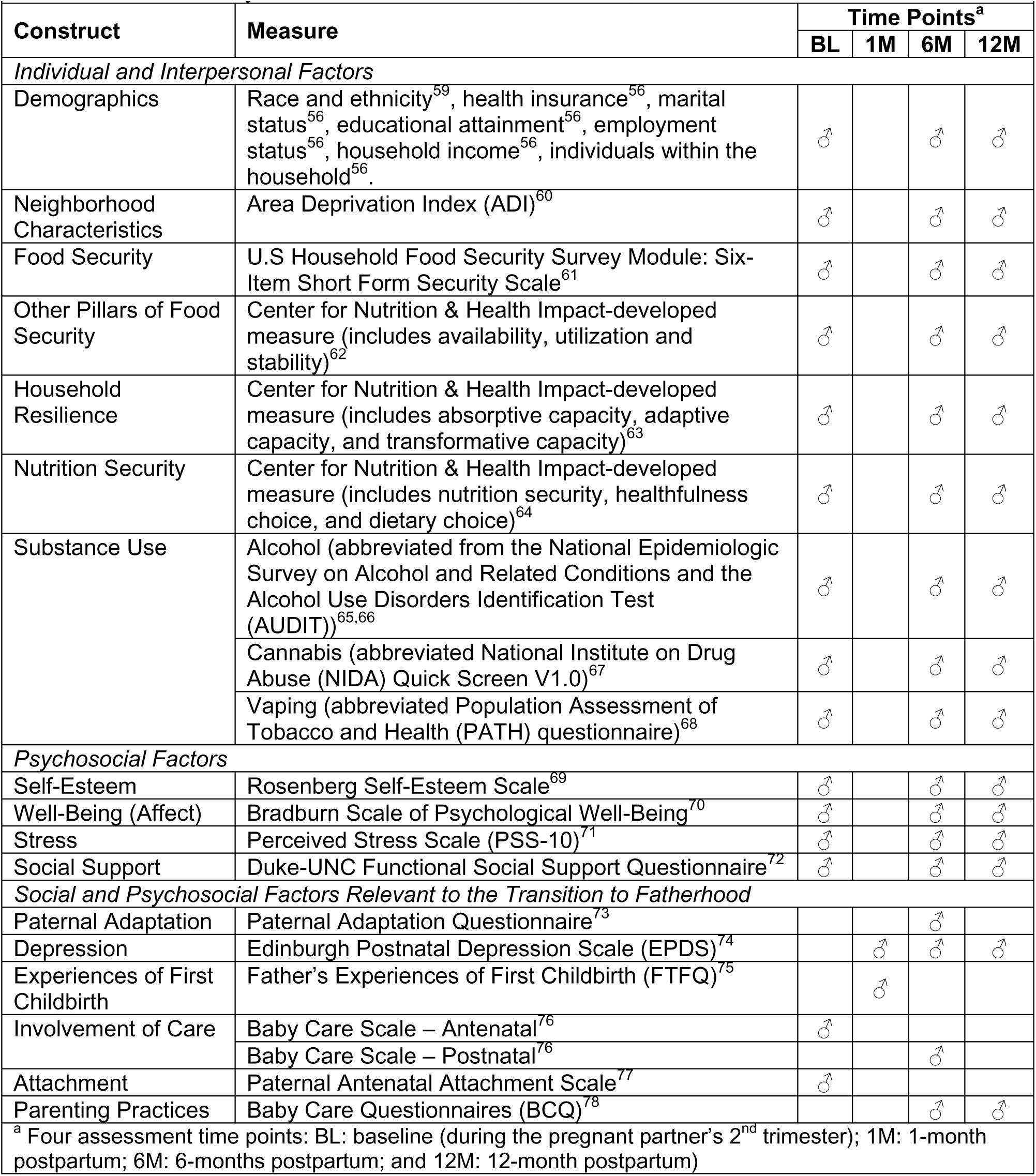
Dad Bod Study Questionnaire Constructs, Measures, and Assessment Schedule.

| Table 2. Dad Bod Study Questionnaire Constructs, Measures, and Assessment Schedule |  |  |  |  |  |
| --- | --- | --- | --- | --- | --- |
| Construct | Measure | Time Points<sup>a</sup> |  |  |  |
|  |  | BL | 1M | 6M | 12M |
| Individual and Interpersonal Factors |  |  |  |  |  |
| Demographics | Race and ethnicity<sup>59</sup>, health insurance<sup>56</sup>, marital status<sup>56</sup>, educational attainment<sup>56</sup>, employment status<sup>56</sup>, household income<sup>56</sup>, individuals within the household<sup>56</sup>. | ♂ |  | ♂ | ♂ |
| Neighborhood Characteristics | Area Deprivation Index (ADI)<sup>60</sup> | ♂ |  | ♂ | ♂ |
| Food Security | U.S Household Food Security Survey Module: Six-Item Short Form Security Scale<sup>61</sup> | ♂ |  | ♂ | ♂ |
| Other Pillars of Food Security | Center for Nutrition & Health Impact-developed measure (includes availability, utilization and stability)<sup>62</sup> | ♂ |  | ♂ | ♂ |
| Household Resilience | Center for Nutrition & Health Impact-developed measure (includes absorptive capacity, adaptive capacity, and transformative capacity)<sup>63</sup> | ♂ |  | ♂ | ♂ |
| Nutrition Security | Center for Nutrition & Health Impact-developed measure (includes nutrition security, healthfulness choice, and dietary choice)<sup>64</sup> | ♂ |  | ♂ | ♂ |
| Substance Use | Alcohol (abbreviated from the National Epidemiologic Survey on Alcohol and Related Conditions and the Alcohol Use Disorders Identification Test (AUDIT))<sup>65,66</sup> | ♂ |  | ♂ | ♂ |
|  | Cannabis (abbreviated National Institute on Drug Abuse (NIDA) Quick Screen V1.0)<sup>67</sup> | ♂ |  | ♂ | ♂ |
|  | Vaping (abbreviated Population Assessment of Tobacco and Health (PATH) questionnaire)<sup>68</sup> | ♂ |  | ♂ | ♂ |
| Psychosocial Factors |  |  |  |  |  |
| Self-Esteem | Rosenberg Self-Esteem Scale<sup>69</sup> | ♂ |  | ♂ | ♂ |
| Well-Being (Affect) | Bradburn Scale of Psychological Well-Being<sup>70</sup> | ♂ |  | ♂ | ♂ |
| Stress | Perceived Stress Scale (PSS-10)<sup>71</sup> | ♂ |  | ♂ | ♂ |
| Social Support | Duke-UNC Functional Social Support Questionnaire<sup>72</sup> | ♂ |  | ♂ | ♂ |
| Social and Psychosocial Factors Relevant to the Transition to Fatherhood |  |  |  |  |  |
| Paternal Adaptation | Paternal Adaptation Questionnaire<sup>73</sup> |  |  | ♂ |  |
| Depression | Edinburgh Postnatal Depression Scale (EPDS)<sup>74</sup> |  | ♂ | ♂ | ♂ |
| Experiences of First Childbirth | Father's Experiences of First Childbirth (FTFQ)<sup>75</sup> |  | ♂ |  |  |
| Involvement of Care | Baby Care Scale – Antenatal<sup>76</sup> | ♂ |  |  |  |
|  | Baby Care Scale – Postnatal<sup>76</sup> |  |  | ♂ |  |
| Attachment | Paternal Antenatal Attachment Scale<sup>77</sup> | ♂ |  |  |  |
| Parenting Practices | Baby Care Questionnaires (BCQ)<sup>78</sup> |  |  | ♂ | ♂ |
| <sup>a</sup> Four assessment time points: BL: baseline (during the pregnant partner's 2<sup>nd</sup> trimester); 1M: 1-month postpartum; 6M: 6-months postpartum; and 12M: 12-month postpartum) |  |  |  |  |  |

### Participant Incentives

Participants will be compensated for participation in the study at each timepoint with a $40 gift card (up to $160 total for completion of all timepoints). Compensation is one method being used to help maximize high retention rates throughout the study. As research volunteers, participants can stop taking part in the study at any point during the study period. In that scenario, participants will be compensated based on the study timepoints they have completed. Following the study’s completion, participants will be allowed to keep the study-provided weight scale and blood pressure cuff.

### Data Management

An electronic software program (RedCap software) will be used to enter and store data and will only be accessible to the authorized members of the research team^38^. All data will be stored in a de-identified format, with personal identifiers removed and replaced with unique study IDs to protect participant privacy.

### Sample Size

Sample size was based on the power to detect the trend regression coefficient in simulation of the Generalized Estimating Equation model (Aim 2). The power to detect a non-null moderate trend effect (−0.05 points in CVH per month^39^) with a sample size of 125 first-time fathers is 0.87 (see **Table 4**).

**Table 4.**
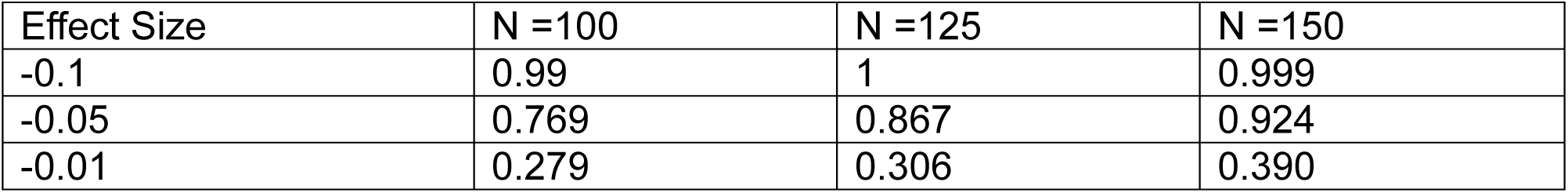
Power to Detect a Non-Null Trend Effect for Various Sample Size and Proposed Interaction Effect Sizes.

| Effect Size | N =100 | N =125 | N =150 |
| --- | --- | --- | --- |
| -0.1 | 0.99 | 1 | 0.999 |
| -0.05 | 0.769 | 0.867 | 0.924 |
| -0.01 | 0.279 | 0.306 | 0.390 |

## Statistical Analysis

For all analyses, baseline data will be described using standard descriptive statistics including frequency distributions, means and SDs, and medians and interquartile ranges. The primary outcome is change in Life’s Essential 8 total score. Analysis plans specific to reach of the three research aims are described in detail below.

### Aim 1 Analysis

Data will be split into training and testing sets with 80% of the sample allocated to training data and the remaining 20% left for testing data. Descriptive statistics of the baseline covariates (means and standard deviations) will be generated to check whether marginal distributions of predictive variables between training and test sets are relatively similar. Should this not be the case, we will regenerate the assignment of observations to the training set until distributions are similar. We will use XGBoost regression as the predictive algorithm with Mean Squared Error (MSE) as the primary loss function^40^. We select model parameters using grid search over the number of splits, training speed, observation subsampling rate, and feature sampling rate optimizing the 5-fold cross-validation MSE. Once the final model parameters are chosen, we will train the model a final time using all of the training data, after which we will report the predictive metrics on the testing data Once the final model is chosen, SHAP scores (Shapley Additive exPlanations) will be computed^41^. For each covariate, Mean Absolute SHAP will be computed to rank each predictor’s importance toward predicting 1-year post-partum CVH. Among predictors with a Mean Absolute SHAP >5 (meaning, on average, the predictor influences the final prediction at least 5 points on the Life’s Essential 8 composite score), we will plot each observation SHAP versus the predictor value. This will give a more in-depth view of how predictor values influence model outputs. Mean absolute SHAP scores and plots will be reported and serve as hypothesis generation tools for future study of targeted interventions aimed at specific population subgroups of young adult men.

### Aim 2 Analysis

Generalized Estimating Equations will be used to model the trajectory of Life’s Essential 8 composite score among first time fathers and will be based on the model described below. Let *μ*_*ij*_ be the mean LE8 composite score of a participant *i* at observation *j*.

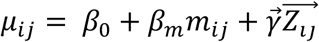

Where *m*_*ij*_ is the number of months to/since birth on observation *j* for participant *i*. *Z*_*ij*_ is the set of confounding predictors for CVH trajectories. We assume an AR (1) correlation structure between successive measurements on the same participant, under the assumption that CVH measurements taken closer together in time will be more correlated than those taken further apart. Based on the group trends observed, we will consider the use of interaction terms between *m*_*ij*_ and a grouping variable that can explain differences in trend.

There are 2 main parameters of interest: *β*_*m*_: comparing two subpopulations of first-time fathers who are similar with respect to 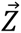 and differing in time till/since birth by 1 month, *β*_*m*_ is the mean difference in Life’s Essential 8 composite score. Parameters will be estimated using a GEE, and the corresponding estimates and 95% sandwich-based confidence intervals will be reported ^42^.

### Aim 3 Analysis

Throughout the study, we will collect data that can be analyzed to identify best practices for recruitment, retention, and remote data collection among first-time fathers for future research. After completing the fourth data collection timepoint, participants will be invited to complete a study feedback questionnaire^43^. Additionally, they will have the option to participate in an exit interview with a study coordinator to discuss their experiences in the study. These data will be reported with exploratory summary statistics.

#### Missing Data

We expect there to be missing data, which will naturally affect the inference in all 3 aims. We will use information from staff feedback and participant study feedback questionnaire to inform the appropriate missingness paradigm. If it is reasonable to assume that missing observations are completely unrelated to cardiovascular health, we will proceed with a complete case analysis for all aims. If the missing observations are related to our observed quantities, we will implement a multiple imputation strategy and amend the inference (p-values, confidence intervals, mean absolute SHAP) to account for the uncertainty in the imputation procedure. If there is non-ignorable missingness, we will present our complete case results with an accompanying worst-case sensitivity analysis over reasonable ranges of what could have been observed.

## Discussion

The Dad Bod Study is a prospective, longitudinal and observational study of young men who are prospective first-time fathers. Research during the perinatal period has predominantly focused on maternal and child health^44,45^, often overlooking the significant biological, social, and psychosocial changes that may be experienced by fathers (**Figure 1**). This bias has left a substantial gap in our understanding of paternal health during this transformative life stage. A better understanding of paternal CVH during the perinatal period is critical to meet new fathers’ needs during this significant life transition. This understanding has implications for both paternal health and the overall wellbeing of the mother-father-infant triad^11^.

The Dad Bod Study brings several innovations in its study design and builds on prior studies examining the impact of fatherhood on men’s health. This study’s longitudinal design, spanning from mid-pregnancy through the first year of fatherhood, allows for a comprehensive assessment of CVH trajectories, capturing both acute and long-term changes associated with new paternal roles and responsibilities. By utilizing the American Heart Association’s Life’s Essential 8 metrics, the study provides a standardized and comprehensive evaluation of paternal CVH, encompassing both behavioral and biological factors. This is compared to other widely used risk assessment tools such as the Pooled Cohort ASCVD Risk Equations^46^ or the more recent American Heart Association Predicting Risk of CVD EVENTs (PREVENT) equations^47^. However, while this study includes several objective metrics of CVH (e.g., dried blood spot for LDL cholesterol), several Life’s Essential 8 metrics are collected from validated self-report measures (e.g., sleep via the Pittsburgh Sleep Quality). This was largely done for feasibility and cost; however, future studies could use objective measures when possible (e.g., accelerometer for measurement of physical activity and sleep). Another notable strength of the study lies in its utilization of comprehensive questionnaires to assess a wide range of social and psychosocial constructs. The data derived from these questionnaires are crucial, as they provide essential context for understanding the factors that influence the potential for improving or maintaining CVH^37^. Lastly, the study’s remote data collection protocol was specifically designed to minimize participant burden by enabling fathers to complete all study components from their home environment. This approach acknowledges the time constraints often experienced by new fathers and may potentially enhance recruitment and retention rates, as well as data quality, by accommodating the participants’ busy schedules and reducing logistical barriers to participation.

This study has limitations that also warrant discussion. First, participants may modify aspects of their behaviors in response to participation in the study (i.e., Hawthorne effect^48^), which may introduce bias into the collection of self-reported health metrics. Additionally, a potential limitation is participation bias, wherein individuals who are already health-conscious may be more likely to participate, potentially affecting the generalizability of the results. However, this study utilizes validated, widely used measures, which can account for some level of potential self-report bias. Generalizability may also be impacted by restricting the sample to English speakers^49^.

In Aim 1 we seek to build a machine learning predictive model for estimating 1-year post-partum CVH from baseline features. Sample size limitations likely inhibit building a truly generalizable model; however, it will be adequate for hypotheses generation for discriminating features of first-time fathers who will decline in CVH. In Aim 2, as our sample is purely composed of first-time fathers, it is difficult to discriminate with precision whether the change in CVH health among first time fathers is due to the effect of simply aging, or whether the birth of their first child resulted in some accelerative effect on CVH. While this study provides critical preliminary data on the transition to fatherhood, in a subsequent study, it would be useful to recruit both first-time fathers and age-matched non-expecting men and follow them over similar amounts of time. Following baseline data collection, this study requires follow-up at three postpartum timepoints. While data collection strategies that minimize participant burden are used, there may be incomplete data among men who are lost to follow-up. Multiple reminder strategies and participant incentives will be implemented across the duration of the study to minimize potential participant attrition. Due to the remote data collection methods^50^, which decrease participant burden, anticipated participant attrition is expected to be low. Any additional challenges to recruitment and retention (Aim 3) will provide important learning opportunities that would strengthen a future study’s design and execution.

## Summary

This prospective, longitudinal and observational study represents a novel examination of CVH among first-time fathers and how CVH may change as young men face new roles and responsibilities associated with fatherhood. The identification of specific factors that are predictive of CVH allow for hypotheses generation of discriminating features of first-time fathers who will decline in CVH. This in turn could set the stage for public health interventions and clinical applications targeted towards these factors. Additionally, outcomes provide novel insights into CVH behaviors and risk factors that would be most critical to monitor, preserve, and improve among young men.

## Data Availability

Data sharing is not applicable to this protocol manuscript as no datasets were generated or analyzed. After study completion, the anonymized data and material will be made publicly available.

## Acknowledgments

None

## Author Contributions

MJL drafted the manuscript. All authors read and approved the final manuscript.

## Data Sharing

Data sharing is not applicable to this article as no datasets were generated or analyzed. After study completion, the anonymized data and material will be made publicly available.

## Non-Standard Abbreviation and Acronyms

CE Studio: Community Engagement Studio
CVD: Cardiovascular Disease
CVH: Cardiovascular Health
SHAP: Shapley Additive exPlanations

